# INOVASIA Study: A Randomized Open Controlled Trial to Evaluate Prophylactic Pravastatin in the Prevention of Preeclampsia and its Effects on sFlt1/PLGF Levels in Pregnant Women at High Risk of Developing Preeclampsia

**DOI:** 10.1101/2021.06.19.21259184

**Authors:** Muhammad Ilham Aldika Akbar, Angelina Yosediputra, Raditya Eri Pratama, Nur Lailatul Fadhilah, Sulistyowati, Fariska Zata Amani, Ernawati, Erry Gumilar Dachlan, Muhammad Dikman Angsar, Gus Dekker

## Abstract

**Objectives:** The study aim is to evaluate the effect of pravastatin to prevent preeclampsia (PE) in pregnant women at a high risk of developing preeclampsia and the maternal and perinatal outcomes and the sFlt1/PLGF ratio in the Surabaya cohort of the INOVASIA trial.

**Setting:** This study involved 2 academic hospital (a tertiary and secondary center) in Surabaya, Indonesia.

**Participants:** Pregnant women at a high risk of developing PE were recruited and randomized into an intervention group (40) and a control group (40). The inclusion criteria consisted of pregnant women with positive clinical risk factor and abnormal uterine artery doppler examination at 10-20 weeks gestational age.

**Inteventions:** The control group received low dose aspirin (80 mg/day) and calcium (1 g/day), while the intervention group received additional pravastatin (20 mg twice daily) starting from 14-20 weeks gestation until delivery. Research blood samples were collected before the first dose of pravastatin, and just before delivery.

**Primary and Secondary Outcomes:** The primary outcome was the rate of maternal preeclampsia, secondary outcomes included maternal-perinatal outcomes and sFlt-1, PLGF, sFlt-1/PlGF ratio and sEng levels.

**Results:** The rate of preeclampsia was (non-significantly) lower in the pravastatin group compared with the control group (17.5% vs 35%). The pravastatin group also had a (non-significant) lower rate of severe preeclampsia, HELLP syndrome, acute kidney injury and severe hypertension. The rate of (iatrogenic) preterm delivery was significantly lower (p 0.048) in the pravastatin group (n=4) compared with the controls (n=12). Neonates in the pravastatin group had significantly higher birthweights, higher Apgar scores, and lower composite neonatal morbidity and NICU admission rates. All biomarkers show a significant deterioration in the control group compared with non-significant changes in the pravastatin group.

**Conclusions:** Pravastatin holds promise in the secondary prevention of preeclampsia and placenta-mediated adverse perinatal outcomes by improving the anti-angiogenic/pro-angiogenic imbalance.

**Trial Registration:** Clinical Trial Gov (ID: NCT03648970)

**Article Summary:** *Strengths and limitations of this study:*

- The largest randomized clinical trial reporting the effects of pravastatin in the prevention of preeclampsia in pregnant women at a high risk of developing preeclampsia with maternal preeclampsia as primary endpoint
- Secondary outpoints included perinatal outcomes, and sFlt-1, PlGF, sFlt-1/PlGF ratio, and sEng levels
- Insufficient funding for placebo tables in the setting of Indonesia, a developing country, resulted in the trial design being an open randomized controlled trial
- In order to reduce risk of bias, 2 independent MFM consultants evaluated and verified all abnormal outcomes while being blinded for group allocation of the trial participants

## INTRODUCTION

Preeclampsia complicates 2-8% of pregnant women worldwide and is one the leading causes of maternal and perinatal morbidity and mortality [1–3]. Preeclampsia may lead to maternal complications such as eclampsia, intracranial bleeding, acute renal failure, pulmonary edema, HELLP syndrome, and DIC. The high fetal/neonatal mortality rate is caused by fetal growth restriction, stillbirth, and complications related to mostly iatrogenic prematurity [4–8].

Over the past decades, the pathogenesis of preeclampsia has been studied extensively. While many theories have been proposed, it is now accepted by most researchers that syncytiotrophoblast (STB) stress is the fundamental pathway leading to the maternal syndrome [6,9,10]. In early-onset preeclampsia, superficial endovascular cytotrophoblast (CTB) invasion in the spiral arteries leading to ischaemia/reperfusion and inflammatory injuries appears to be the most important pathway leading to STB stress. While in the much more common late-onset preeclampsia, the STB stress appears to be primarily related to maternal constitutional and lifestyle related factors [5,6,8–13]. STB stress will lead to the release of various pro-inflammatory and anti-angiogenic factors in the maternal systemic circulation, causing the characteristic maternal endothelial cell dysfunction [6,11]. Many recent studies have demonstrated that an imbalance between pro-angiogenic (VEGF and PLGF) and anti-angiogenic (sFlt-1, s-Eng) factors plays a pivotal role in the pathophysiology of preeclampsia [11,12]. Both anti-angiogenic markers are known to increase significantly 8-12 weeks before the onset of preeclampsia [12–15].

Until now, the only definitive treatment for preeclampsia is delivery of the baby and placenta [2,3,16,17]. Recent research has focused on finding new ‘curative’ drugs for preeclampsia. Ahmed et al were the first to propose statins as a new therapeutic strategy [18– 20]. According to Ahmed et al, decreased activity of Heme oxygenase 1 (HO-1) in early pregnancy may trigger a negative cascade of events such as oxidative stress, inflammation, and elevated sFlt-1 and sEng levels [21–23]. HO-1 is a rate limiting enzyme with as primary duty the breakdown of heme into biliverdin, free iron, and carbon monoxide (CO) in the cellular endoplasmic reticulum[22]. Statins are a group of cholesterol lowering drugs used mainly in the treatment of hypercholesterolemia. The rationale to use statin as a drug for preeclampsia is based on animal studies showing that pravastatin (3-hydroxy-3-methylglutaryl coenzyme-A-reductase inhibitor) has a protective role at the uteroplacental interface and on vascular cells [18]. Pravastatin may protect endothelial cells, by inducing the expression of HO-1 and as such inhibiting cytokine mediated release of the anti-angiogenic factors sFlt-1 and sEng [4,19,20,22,24–27]. Decreased levels of sFlt-1 and sEng may increase free PlGF and VEGF in the circulation. Correcting the imbalance may eventually resolve the maternal endothelial cell dysfunction. Statins have pleiotropic effects which may, in theory, be beneficial in preventing preeclampsia including: immunomodulatory and anti-inflammatory effects, and reduction in free oxygen radical formation and smooth muscle cell proliferation [4]. We recently demonstrated that pravastatin reduces levels of pro-inflammatory cytokines and endothelial activation markers in patients at risk of developing preeclampsia[28].

There are relatively limited data on the safety of pravastatin during pregnancy. However, so far animal and human data have not found an increased risk of congenital anomalies in infants after statin use in pregnant women [29–31]. A systematic review and meta-analysis about pregnancy outcome following first trimester exposure to statins (reviewing all studies until 2013), also came to reassuring conclusions: with a total of more than 800 patients (from 6 studies), the results showed no increased risk of birth defects in the statin-exposed pregnancies compared with the control subjects (RR 1.15; 95% CI 0.75 to 1.76) [32]. In addition, pravastatin is hydrophilic, levels in the fetoplacental unit are therefore quite low compared with other statins [33–37].

Ahmed et al, started the first pilot study (STAMP trial; pravastatin to ameliorate early onset preeclampsia) in the UK [38]. The STAMP trial failed to show any decrease in sFlt-1 level in early onset preeclampsia women treated with pravastatin [38]. However, this study was performed in already severely sick preterm preeclamptic women which may have limited the efficacy of the treatment. Rather than trying to treat the disease, it might be better to prevent preeclampsia from the beginning to reduce the risk of adverse maternal-neonatal outcomes. In 2017, we started the multicenter INOVASIA study (*Indonesia Pravastatin to Prevent Preeclampsia study*; Clinical trial Gov ID: NCT03648970) involving 3 central tertiary hospitals in Indonesia. The Surabaya center also measured a series of relevant biomarkers in addition to the clinical outcomes. The aim of this paper is to present the effects of pravastatin on sFlt-1, PlGF, and sEng levels, and the sFlt-1/PlGF ratio in conjunction with the maternal and perinatal outcome in the Surabaya cohort in pregnant women at a high risk of developing preeclampsia.

## MATERIAL & METHODS

### Sample Recruitment, Definition, Inclusion and Exclusion criteria

The Surabaya arm of the INOVASIA study (2017-2020) was performed at Universitas Airlangga Academic Hospital and Dr. Soetomo General Academic Hospital. The current research paper is part of the clinical multicenter INOVASIA trial (Indonesia Pravastatin to Prevent Preeclampsia Study) which has been registered in Clinical Trial Gov (ID: NCT03648970). INOVASIA is an ongoing multicenter randomized controlled trial in Indonesia with the aim to evaluate the effects of pravastatin (in addition to low dose Aspirin and calcium) on maternal and perinatal outcomes in pregnant women at a high risk of developing preeclampsia women compared with a control group. The INOVASIA study was approved by the Ethical and Law Committee of Universitas Airlangga Academic Hospital (122/KEH/2017) and The Ethical Committee in Health Research Dr. Soetomo General Academic Hospital Surabaya (427/Panke.KKE/VII/2017). Only the Surabaya centers had the capacity to include the measurement of a series of relevant biomarkers.

Women deemed to be at a high risk of developing preeclampsia women were identified at 10-20 week’s gestation using a combination of clinical and ultrasound criteria. The clinical risk factors included: history of preeclampsia, family history of preeclampsia, obesity, diabetes, chronic hypertension, vascular collagen disease, nulliparity, maternal age > 40 years, mean arterial pressure (MAP) > 90 mm Hg, and abnormal uterine artery Doppler velocimetry. Uterine artery Doppler velocimetry examination was performed based on standard protocol by the Maternal-Fetal Medicine Foundation [37,38]. The criteria of abnormal Doppler velocimetry index were based on the pulsatility index (PI), or resistance index (RI), or presence of early diastolic notching. In the first trimester ultrasound: PI > 95th percentile and in the second trimester: early bilateral diastolic notching or RI > 0.58 were criteria for abnormal Doppler of uterine artery [39–41].

The aim was to select women with a preeclampsia risk of at least 20% as based on the presence of minimally 2 independent clinical risk factors. Exclusion criteria included: current statin use, known statin hypersensitivity, acute liver disease, or participation in any other clinical trial. Thirty-five patients (49.3%) were recruited at 11-14 weeks, the remaining 41.7% between 14-20 week’s gestation (table 1). Preeclampsia was diagnosed using the International Society for The Study of Hypertension in Pregnancy (ISSHP) criteria (2018) [39]. Chronic hypertension was defined as high blood pressure (≥ 140/90) before pregnancy or before 20 weeks gestation, or being medicated because of known chronic hypertension[39]. Obesity was diagnosed based on measurement of body mass index (BMI) value at recruitment ≥ 30 kg/m^2^ [40,41]. Patients with history of systemic lupus erythematosus, rheumatoid arthritis, or scleroderma were defined as having a collagen vascular disease.

**Table 1.** General Maternal Characteristics and Preeclampsia Risk Factors.

| Maternal Characteristics | Control<br>n = 40 | Pravastatin<br>n = 40 | p value |
| --- | --- | --- | --- |
| Gestational Age at recruitment | 15 (14-20) | 16.5 (14-20) | 0.304<sup>c</sup> |
| Maternal Age | 29.75 ± 5.93 | 31.98 ± 5.72 | 0.597<sup>a</sup> |
| Parity |  |  |  |
| Nullipara | 16 (40%) | 9 (22.5%) | 0.147<sup>b</sup> |
| Multipara | 24 (60%) | 31 (77.5%) |  |
| Gravidity | 2 (1-5) | 2 (1-7) | 0.168<sup>c</sup> |
| BMI (kg/m<sup>2</sup>) | 25.41 ± 6.78 | 28.33 ± 5.72 | 0.415<sup>a</sup> |
| **Preeclampsia Risk Factors** |  |  |  |
| Systolic blood pressure at initial visit | 116.38 ± 15.06 | 122.65 ± 11.16 | 0.037<sup>a</sup> |
| Diastolic blood pressure at initial visit | 80 ± 11.25 | 82.03 ± 9.55 | 0.388<sup>a</sup> |
| Mean arterial blood pressure at initial visit | 92.12 ± 11.75 | 95.57 ± 9.29 | 0.150<sup>a</sup> |
| MAP ≥ 90 mmHg at initial visit | 26 (65%) | 31 (77.5%) | 1.525<sup>b</sup> |
| Obesity (BMI ≥ 30 kg/m<sup>2</sup>) | 11 (27.5%) | 14 (35%) | 0.523<sup>b</sup> |
| History of iatrogenic preterm birth | 6 (15%) | 1 (2.5%) | 0.108<sup>b</sup> |
| History of stillbirth | 2 (5%) | 4 (10%) | 0.675<sup>b</sup> |
| History of chronic hypertension | 8 (20%) | 7 (17.5%) | 1.000<sup>b</sup> |
| History of diabetes mellitus | 2 (5%) | 2 (5%) | 1.000<sup>b</sup> |
| History of renal disease | 2 (5%) | 0 | 0.494<sup>b</sup> |
| History of vascular collagen disease | 4 (10%) | 1 (2.5%) | 0.359<sup>b</sup> |
| History of preeclampsia | 6 (15%) | 5 (12.5%) | 1.000<sup>b</sup> |
| History of gestational diabetes mellitus | 2 (5%) | 0 | 0.494<sup>b</sup> |
| Family history of preeclampsia | 1 (2.5%) | 2 (5%) | 1.000<sup>b</sup> |
| Family history of chronic hypertension | 4 (10%) | 3 (7.5%) | 1.000<sup>b</sup> |
| **Uterine artery Doppler velocimetry** |  |  |  |
| PI Uterine Artery | 1.39 ± 0.64 | 1.29 ± 0.65 | 0.561 |
| RI Uterine Artery | 0.63 ± 0.19 | 0.65 ± 0.08 | 0.756 |
| S/D Uterine Artery | 2.72 ± 1.76 | 2.58 ± 1.72 | 0.778 |

<sup>a</sup>Independent T-test; <sup>b</sup>Chi Square test; <sup>c</sup>Mann Whitney test; Red color: $p < 0.05$ ; NA: Not Available

### Intervention

Patients meeting the inclusion criteria were randomized into the control or pravastatin group using computer randomization. The control group received the standard low dose Aspirin (80 mg) and calcium (1000 mg) orally. The pravastatin group received pravastatin 20 mg tablets twice daily in addition to the aforementioned standard drugs. Low dose Aspirin and calcium were started from < 20 week’s gestation until 36 week or until preeclampsia symptoms developed. Pravastatin was started directly following recruitment of patients (< 20 weeks) and continued until delivery. Patients were followed until delivery; post-delivery the various clinical outcome variables were collected (figure 1).

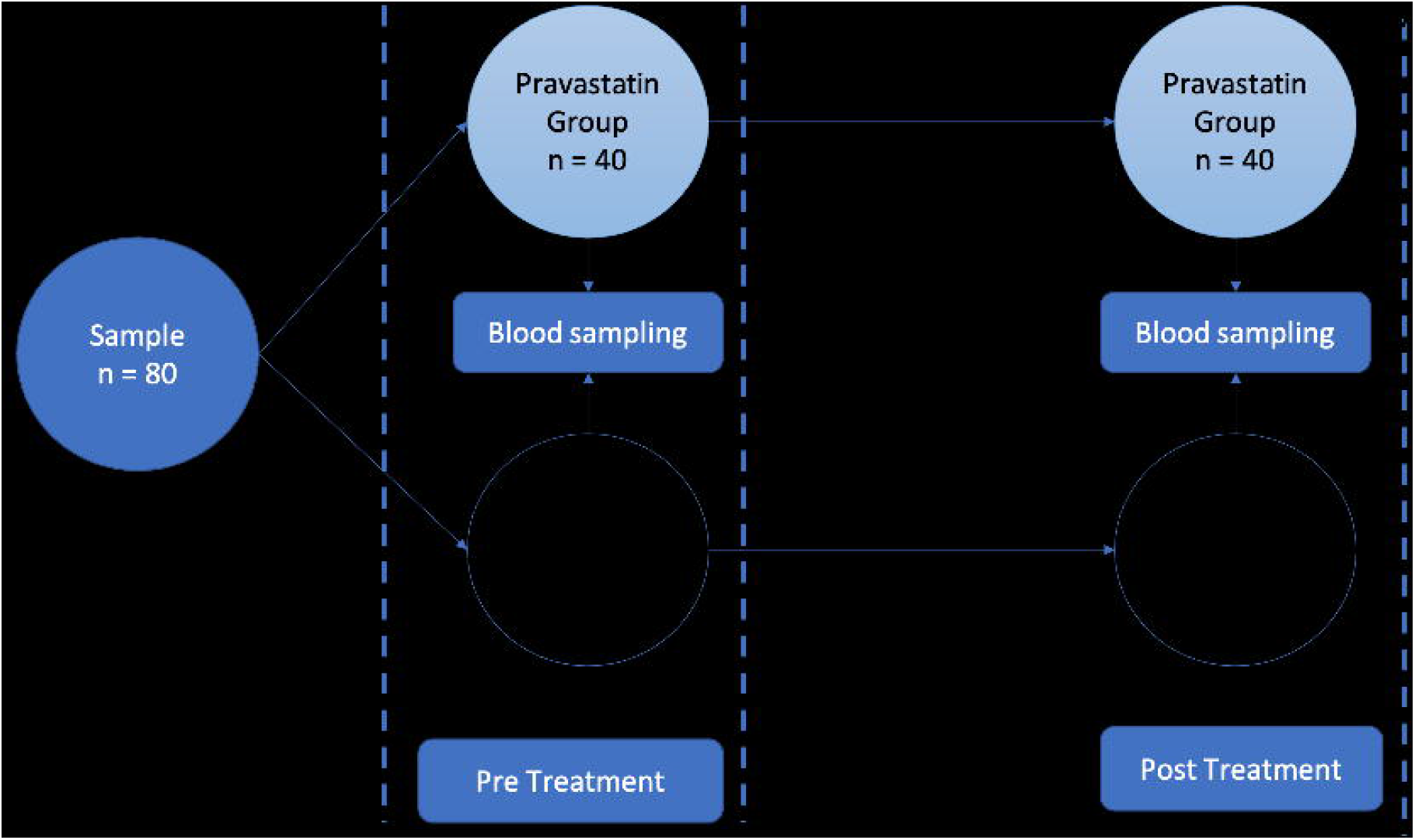

Research maternal blood samples (10 ml) were taken twice: 1. Immediately after recruitment, prior to the start of pravastatin (pre-treatment); 2. During early labour or before induction of labour/caesarean section (post-treatment) (figure1). After initial processing (centrifugation 1000 RPM for 15 minutes (< 60 min)), serum samples were stored in -80°C freezer until all participants had completed the trial. All the samples were processed and analyzed simultaneously.

The primary outcome of the study was occurrence of preeclampsia. Secondary endpoints included : preeclampsia with severe features, maternal mortality, preterm delivery < 37 and < 34 weeks gestation, maternal complications (pulmonary oedema, eclampsia, acute kidney injury, HELLP syndrome, severe hypertension (≥ 180/110 mmHg), disseminated intravascular coagulation), mode of delivery, neonatal birthweight and length, Apgar score 1^st^ minute and 5^th^ minutes, stillbirth, neonatal death, Small Gestational Age (SGA) (birthweight < 5^th^ centile), respiratory distress syndrome, Neonatal Intensive Care Unit (NICU) admission, ventilator use and composite neonatal morbidities (any occurrence of fetal-neonatal morbidities or mortalities previously mentioned). All adverse pregnancy outcomes were verified by 2 independent reviewers, being unaware of group allocation of the individual trial participants. Laboratory outcomes included maternal serum level of sFlt-1, PlGF, sFlt-1/PlGF ratio and sEng before and after treatment.

### Biomolecular Process and Analysis

Serum biomarkers were analyzed using specific reagen-kits: sFlt-1 (Quantikine Human Soluble VEGF R1/Flt-1 Immunoassay, R&D Systems, Inc, Minneapolis, USA), PlGF (Quantikine Human PlGF Immunoassay ELISA kit, R&D Systems, Inc, Minneapolis, USA), sEng (Quantikine ELISA Human Endoglin/CD 105 Immunoassay kit, R&D Systems, Inc, Minneapolis, USA [42]). The sFlt-1, PlGF, and sEng ELISA kits are the same assays previously used in the original Levine et al study [12,43]. In women with preeclampsia, the correlations between ELISA kit (R&D) and Elecsys (Roche) kit for plasma sFlt-1, PlGF, and sFlt-1/PlGF ratio are 0.98, 0.76, and 0.98, respectively [44]. The Human Soluble VEGF R1/Flt-1, Human Endoglin/CD 105 kit measure the sFlt-1, PlGF, and sEng levels using a quantitative sandwich enzyme immunoassay technique. Intra-assay precision of the sFlt-1, PlGF, sEng reagen-kits were (2.6-3.2%; 5.6-7%; 2.8-3%), while the inter-assay precision was (5.5-9.8%; 10.9-11.8%; 6.3-6.7%). All the laboratory process and analysis were performed in PRODIA laboratory, Surabaya.

### Statistical Analysis

The IBM© SPSS© Statistic ver 25 was used to analyze the data. All data were analyzed with descriptive statistics methods to evaluate mean, median, standard deviation, standard error, and interquartile range, where appropriate. The normality of data was analyzed using the Kolmogorov-Smirnov test, because the number of samples > 50. Categorical data were compared using Chi-square test or Fischer exact test based on their requirements. The numerical data in maternal characteristics were compared using Independent sample T-test (data with normal distribution) or Mann-Whitney test (data with abnormal distribution). The pre- and post-test serum marker levels were compared using Paired sample T-test (data with normal distribution) or the Wilcoxon test (data with abnormal distribution). Normally distributed data are presented as mean ± standard deviation, while the abnormally distributed data are presented as median (interquartile range). The required sample size of the multicenter Inovasia study was 240 participants, aiming for a 50% reduction in the pravastatin group with an expected 20% rate of preeclampsia in the control group. No separate sample size calculation was performed for the Surabaya arm of the Inovasia study.

### Patients and Public Involevement

No patients involved.

## RESULTS

Eighty participants were recruited and randomized into 40 patients in the control group and 40 in the pravastatin group. 50 participants were recruited based on a one or more positive clinical risk factors and abnormal uterine artery Doppler, while the other 30 participants were recruited based on 2 or more maternal clinical risk factors only. Maternal risk factors and further characteristics at time of randomization were similar (table 1). The pravastatin group had a slightly higher systolic blood pressure at recruitment (122.65 ±11.16 vs 116.37 ±15.05 mmHg; *p*=0.037). All participants were recruited before 20 weeks gestation; 49% at 11-14 weeks, the remaining 51% between 14-20 week.

Maternal outcomes are presented in table 2. In the control group 14 out of the 40 women (35%) developed preeclampsia, indicating an appropriate selection of high-risk patients, compared with only 7 (17.5%) in the pravastatin group (OR: 2.54; 95% CI: 0.89-7.2). In the pravastatin group, only 2 (5%) of patients developed preeclampsia with severe features versus 6 (15%) in the control group (OR: 3.35; 95% CI: 0.63-17.74). Major maternal complications like HELLP syndrome, acute kidney injury, and severe hypertension did not occur in the pravastatin group compared with 4 patients (10%) in the control group. Preterm delivery rate < 37 weeks occurred in 4 (10 %) in the pravastatin group compared with 12 (30 %) in the control group (OR: 3.86; 95% CI: 1.12-13.26) (table 3). Of these 16 preterm births, 14 were iatrogenic and 2 spontaneous. The pravastatin group also had a lower preterm delivery rate < 34 weeks compared to control group (7.5% vs 12.5%; NS) (table 3). Caesarean section rates were similar, n=20 in the pravastatin group versus n=24 in the control group. Of the 36 vaginal births, 22 followed induction of labour.

**Table 2.** Maternal Outcomes.

| Clinical Parameters | Control<br>n = 40 | Pravastatin<br>n = 40 | *p* value |
| --- | --- | --- | --- |
| Any preeclampsia | 14 (35%) | 7 (17.5%) | 0.126<sup>b</sup> |
| Severity preeclampsia |  |  |  |
| Preeclampsia | 8 (20%) | 5 (12.5%) |  |
| Preeclampsia with severe features | 6 (15%) | 2 (5%) |  |
| Preeclampsia Type |  |  |  |
| Early Onset | 5 (12.5%) | 2 (5%) |  |
| Late Onset | 9 (22.5%) | 5 (12.5%) |  |
| HELLP Syndrome | 1 (2.5%) | 0 | 1.000<sup>b</sup> |
| Acute Kidney Injury | 1 (2.5%) | 0 | 1.000<sup>b</sup> |
| Severe Hypertension | 2 (5%) | 0 | 0.494<sup>b</sup> |
| Mode of Delivery |  |  |  |
| Vaginal | 16 (40%) | 20 (50%) | 0.500<sup>b</sup> |
| Cesarean Section | 24 (60%) | 20 (50%) |  |

<sup>a</sup>Independent T-test; <sup>b</sup>Chi Square test; <sup>c</sup>Mann Whitney test; <sup>d</sup>Kolmogorov-Smirnov; Red color: $p < 0.05$ ; NA: Not Available

**Table 3.** Perinatal Outcomes.

| **Clinical Parameters** | **Control**<br>**n = 40** | **Pravastatin**<br>**n = 40** | **p value** |
| --- | --- | --- | --- |
| Gestational Age at Delivery | 37.5 (24-39) | 38 (33-40) | 0.315<sup>c</sup> |
| Preterm delivery < 37 weeks | 12 (30%) | 4 (10%) | 0.048<sup>b</sup> |
| Preterm delivery < 34 weeks | 5 (12.5%) | 3 (7.5%) | 0.712<sup>b</sup> |
| Birthweight | 2625 ± 872 | 2931 ± 537 | 0.006<sup>a</sup> |
| Neonatal length | 48 (23-53) | 49 (40-54) | 0.407<sup>c</sup> |
| Apgar Score 1st minutes < 7 | 11 (27.5%) | 1 (2.5%) | 0.002<sup>c</sup> |
| Apgar Score 5th minutes < 7 | 5 (12.5) | 0 | 0.021<sup>c</sup> |
| Neonatal death | 1 (2.5 %) | 0 | 1.000<sup>b</sup> |
| SGA < 5<sup>th</sup> centile | 3 (7.5%) | 0 | 0.241<sup>b</sup> |
| Stillbirth | 1 (2.5%) | 0 | 1.000<sup>b</sup> |
| Respiratory distress | 3 (7.5%) | 0 | 0.241<sup>b</sup> |
| Composite Neonatal Morbidities | 8 (20%) | 0 | 0.005<sup>b</sup> |
| NICU Admission | 6 (15%) | 0 | 0.026<sup>b</sup> |
| Ventilator Used | 3 (7.5%) | 0 | 0.241<sup>b</sup> |

<sup>i</sup>Independent T-test; <sup>b</sup>Chi Square test; <sup>c</sup>Mann Whitney test; Red color: $p < 0.05$ ; NA: Not Available

In line with the significantly lower preterm birth rate, birthweights were significantly higher, also 1- and 5 -minute Apgar scores were better, all resulting in significantly lower composite neonatal morbidity in the pravastatin group (table 3). Neonates delivered in the pravastatin group had no neonatal morbidity and/or NICU admission, while the control group had a rate of 20% and 15%, respectively. One stillbirth and one neonatal death occurred in the control group, while there was no perinatal death in the pravastatin group. On neonatal examination, no neonate in the pravastatin group had any structural malformation.

The sFlt-1 levels significantly increased in the control group (pre vs post treatment (median [IQR]): 2303 [1673] vs 3783 [5253] pg/mL; p=0.007), compared with a modest, non-significant increase in the pravastatin group (2119 [1725] vs 2724 [2786] pg/mL; p=0.461) (figure 2). The PlGF level decreased significantly in the control group (213 [316.8] vs 57.9 [262.1]; *p*=0.013), but again only a minor non-significant decrease was seen in the pravastatin group (306.7 [461.5] vs 200.5 [236.3]; *p*=0.098) (figure 2). Accordingly, the sFlt-1/PlGF ratio significantly increased in control group (10.66 [15.28] vs 28.22 [796.3]; *p*=0.000), with virtually no change in pravastatin group (9.2 [24.06] vs 14.42 [40]; *p*=0.128) (figure 2). The sEng levels in the control group increased more than 3-fold (2208.9 [1877.5] vs 7904.8 [3286.9]; p<0.001), while levels even showed a (non-significant) decrease in the pravastatin group (2975.2 [2155.3] vs 2993.9 [1439.7]; *p*=0.266) (figure 2). All biomarker results are summarized in table 4, and figure 2.

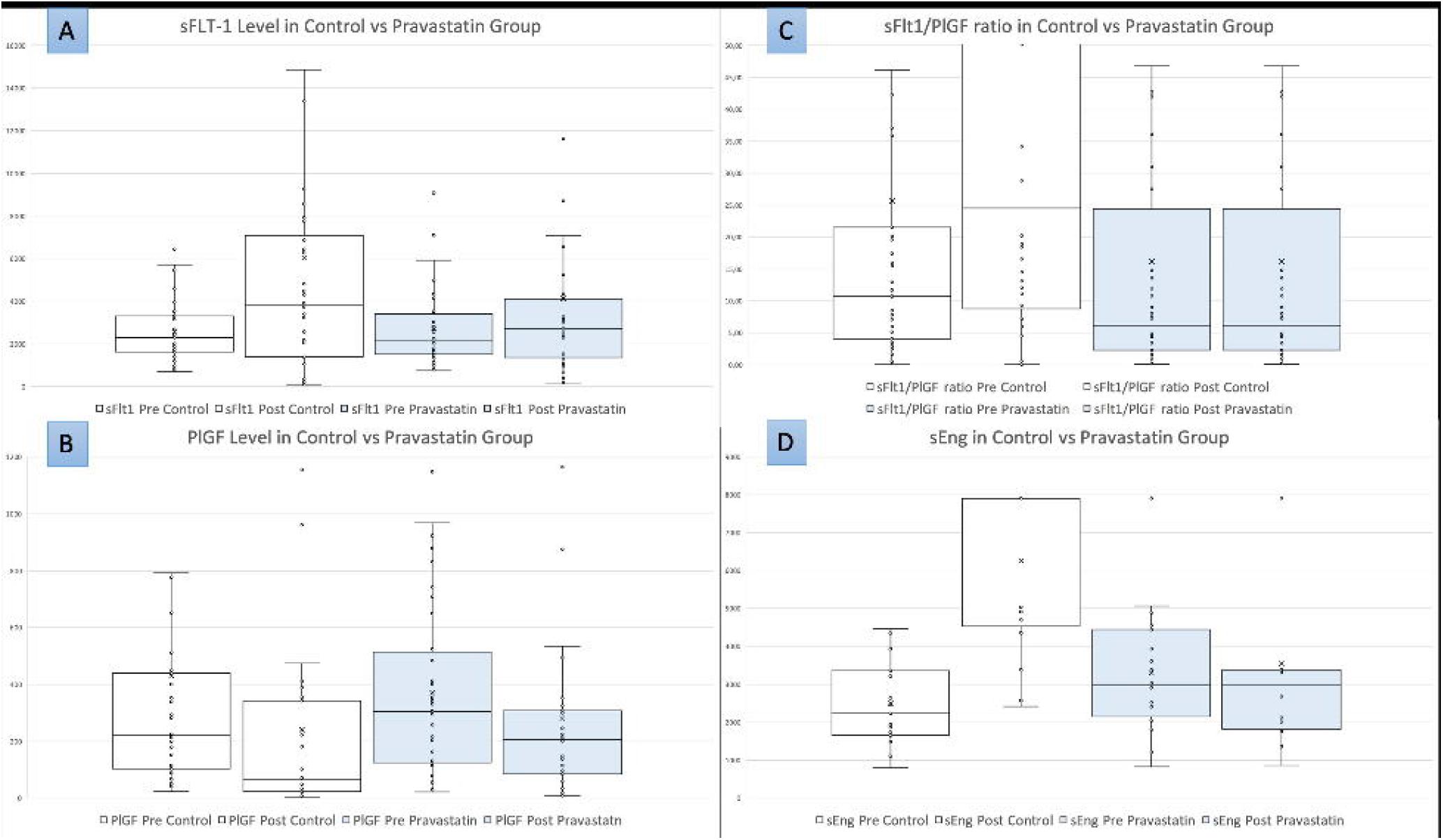

**Table 4.**
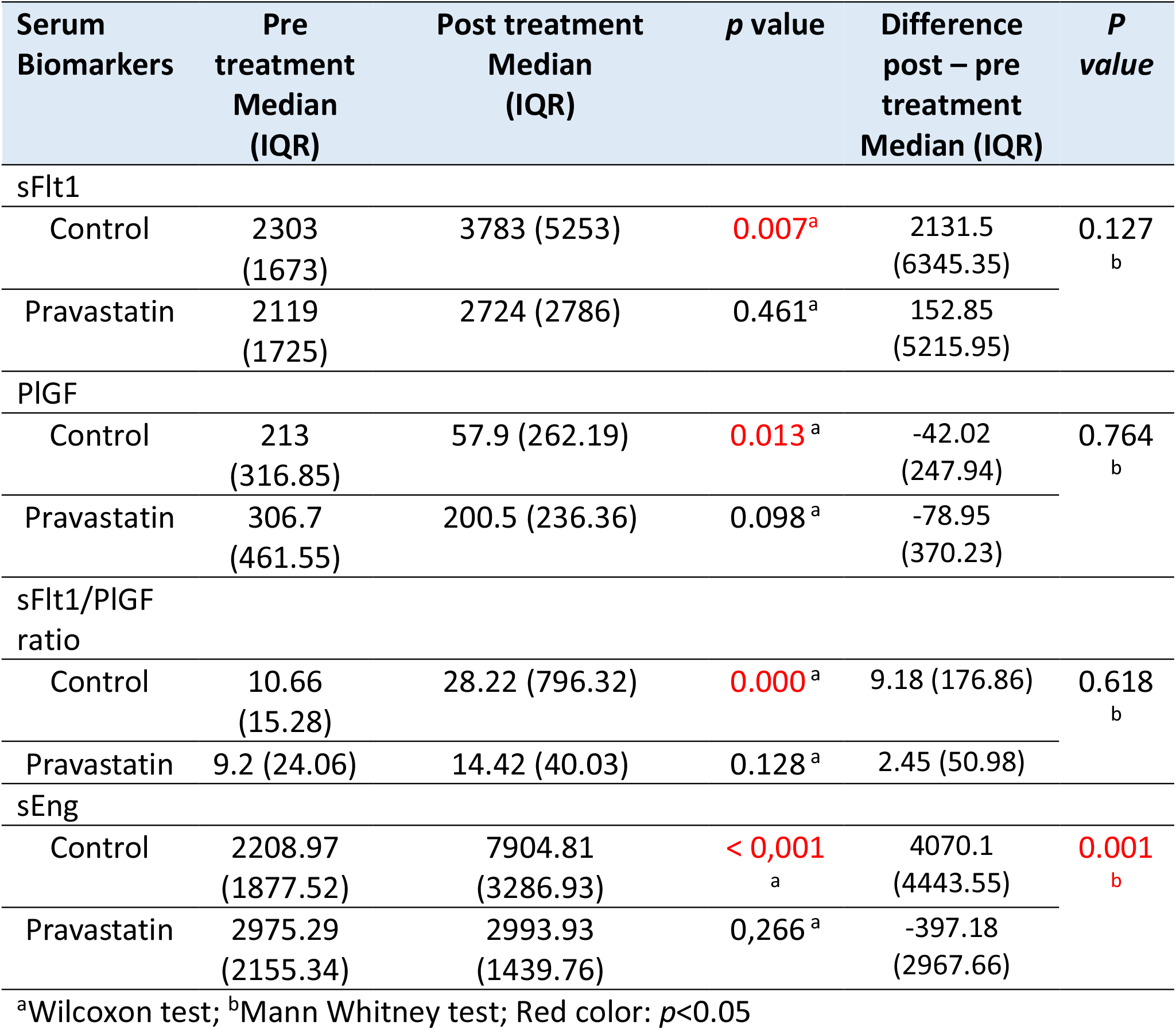
Maternal Serum Biomarkers Level Before and After Treatment.

The serum sFlt-1/PlGF ratios in preeclamptic patients (n=59) versus normal outcomes pregnancies (n=21) were also evaluated. The sFlt-1/PlGF ratio at delivery in preeclamptic patients were (non-significantly) higher compared with the normotensive patients (34.35 [103.20] vs 16.57 [42.17]; *p*=0.322), but with a significantly higher preterm birth rate in the preeclamptic patients (47.6% vs 6.4%; *p*=0.000). The preeclamptic patients had a significantly lower mean gestational age at delivery compared to normotensive patients (34.67 ±3.86 vs 37.46 ±2.259 week; *p*=0.000).

## DISCUSSION

The use of prophylactic pravastatin (on top of low-dose Aspirin and calcium) in women at high risk of developing preeclampsia halved (non-significant) the rate of preeclampsia, and importantly significantly improved the perinatal outcome. The relatively small sample size in the current study, being just the Surabaya arm of the Inovasia study, might explain the lack of statistical significance (p = 0.126) in the reduction of the preeclampsia rate. The rate of preeclampsia with severe features and preeclampsia complications were also markedly lower (non-significant). The use of pravastatin was also associated with an improvement in the sFlt1/PLGF ratio and sEng levels compared with the control group.

Over the past years, pravastatin has been proposed by several researchers as a potentially promising drug to prevent preeclampsia and other placenta mediated disorders based on its pleiotropic potentially beneficial effects on endothelial cells, including anti-oxidant, anti-inflammatory, and antithrombotic effects, and its effect on the angiogenic imbalance [4,22,24,45–52]. After a promising report on a small case series [51], the UK STAMP trial was the first proper trial to evaluate the effect of pravastatin in women with established early-onset preeclampsia [37]. Pravastatin 40 mg daily was given in 30 participants matched with healthy control (n = 32). The outcome of the STAMP trial was that the use of pravastatin did not lead to a reduction in sFlt-1 levels, also maternal blood pressure and time from randomization to delivery between both groups were similar[37]. Following the STAMP trial, Constantine et al reported on a pilot randomized controlled trial [53] in high-risk women, aimed at evaluating the safety and pharmacokinetics of Pravastatin. The rate of preeclampsia was a secondary outcome [53]. The pravastatin group (n=10) had no case of severe preeclampsia compared with 4 cases in the control group (n=10).

This report, on the Surabaya arm of the INOVASIA study, is the first randomized study on the use of prophylactic pravastatin with preeclampsia as primary outcome. The reason for this initial report on the Surabaya arm within this multicenter trial relates to the fact that the Surabaya center was the only center with the capacity to look at various biomarkers and levels of angiogenic and anti-angiogenic factors [28].

Besides the reduction (non-significant) in the preeclampsia rate, the most important finding was the significantly improved perinatal outcome (planned secondary outcome) in the pravastatin group. The rate of preterm delivery (<37 weeks) in pravastatin group is one third compared with control patients. Although not significant, pravastatin may also reduce the risk of preterm delivery < 34 weeks. As a consequence, birthweights in the pravastatin group were significantly higher. The use of pravastatin was also associated with a significantly lower rate of overall perinatal morbidity and NICU admission, with no neonatal morbidities (neonatal death, SGA < 5^th^ centile, stillbirth, RDS, NICU admission, and ventilator use) in the pravastatin group, compared with an overall composite neonatal morbidity of 20% amongst control patients. One neonatal death in the control group related to extreme prematurity following PPROM (24 weeks - 600 g). The stillbirth in the control group occurred in a patient with complicated early-onset preeclampsia (26 weeks, renal failure, uncontrolled severe hypertension [210/110 mmHg]).

The other important pre-specified outcome was the beneficial effect of 40 mg of pravastatin on various serum markers. We recently published on the beneficial effects of pravastatin on various inflammatory cytokines and endothelin levels [28]. The current paper shows that administration of pravastatin started before 20 weeks more or less stabilizes sFlt-1 and PlGF levels, the sFlt-1/PlGF ratio, and sEng levels. While in the control group, we found the expected deterioration of these variables. It is important to emphasize that a degree of STB stress also occurs in uncomplicated pregnancies towards term [6]. Our study design did not allow (financial restrictions) for parallel timing of blood samples, the post-treatment samples were obtained just prior to giving birth. It is therefore important to reflect on the fact that in the pravastatin group only 4 women (10%) gave birth prior 37 weeks compared with 12 (30%) in the control group; if anything, the more advanced gestational age in the pravastatin group would be expected to lead to high sFlt-1 levels, and lower PLGF levels – our findings are quite the opposite. We also note that the sFlt-1/PlGF ratio in preeclampsia group are non-significantly higher compared with normotensive women. The significantly lower gestational age at delivery, and thus lower gestational age at time of blood sampling in the preeclampsia group may explain this lack of statistical significance.

### Strength and Limitation of The Study

This is the largest study reporting on the effects of prophylactic pravastatin in women at high risk of developing preeclampsia. Over the coming year we will be able to present the clinical outcome in the overall INOVASIA study. An obvious limitation is the lack of a placebo group. The reason for this is the very limited funding availability for this research in Indonesia. Obtaining the pravastatin tablets (normally not available in Indonesia used 80% of the research budget, the remaining 20% was used for the bioassays). We do acknowledge the desirability of the double blind RCT design, but the very restricted research funds available and the high cost of placebo tablets made a double-blind approach impossible. Pravastatin is not available in Indonesia, so it would be very unlikely that patients in the control group could have had access to the drug. We also tried to further reduce risk of bias by 2 independent MFM consultants evaluating and verifying the outcomes while being blinded for group allocation of reviewed participants.

## CONCLUSION

Prophylactic pravastatin in pregnant women at a high risk of developing preeclampsia significantly improves perinatal outcome and reduces (non-significant) the risk of preeclampsia. The improved perinatal outcome with prophylactic pravastatin goes hand-in-hand with reduced sFlt-1 and sEng levels and improved sFlt-1/PlGF ratios.

## Data Availability

The data of this study are available from Universitas Airlangga Hospital and Dr. Soetomo General Hospital but restrictions apply to the availability of these data and so are not publicly available. Data are however available from the authors upon reasonable request and with permission of Universitas Airlangga Hospital and Dr. Soetomo General Hospital.

## Authors Contribution

1. Muhammad Ilham Aldika Akbar: Concept and design of study, study implementation and execution, interpretation and analysis of the data, drafting and revising the various drafts and the final version of this manuscript, responsible for submission and communication with the journal.
2. Angelina Yosediputra: Patient selection and recruitment, and maternal-perinatal data collection
3. Raditya Eri Pratama: Patient selection and recruitment, and maternal-perinatal data collection
4. Nur Lailatul Fadhilah: Patient selection and recruitment, and maternal-perinatal data collection
5. Sulistyowati: Patient selection and recruitment, and maternal-perinatal data collection
6. Fariska Zata Amani: Patient selection and recruitment, and maternal-perinatal data collection
7. Ernawati: Drafting and revising the various drafts and the final version of this manuscript
8. Erry Gumilar Dachlan: Concept and design of study, drafting and revising the various drafts and the final version of this manuscript
9. Muhammad Dikman Angsar: Concept and design of study, drafting and revising the various drafts and the final version of this manuscript
10. Gus Dekker: Concept and design of study, interpretation and analysis of the data, drafting and revising the various drafts and the final version of this manuscript

## ACKNOWLEDGEMENT

We thank Prof. Johanes Cornelius Mose MD, PhD, Nadir Abdullah MD, Aditiawarman MD, PhD, Agus Sulistyono MD, PhD, Budi Wicaksono MD, Khanisyah Erza Gumilar MD, Manggala Pasca Wardhana MD, Rozi Aditya Aryananda MD, and Nareswari Cininta MD for their valuable input, advices, and suggestions to complete this research. We thank Prof. Sri Sulistyowati MD, PhD and Fransiscus Oktavius Hari Prasetyadi, MD as the independent reviewers in this study.

